# Pediatric Influenza Vaccine Effectiveness Against Emergency Visits and Hospitalization in Canada: 2024-2025 and 2025-2026 seasons

**DOI:** 10.64898/2026.11.03.26367131

**Authors:** Jeffrey C. Kwong, David M. Goldfarb, Maala Bhatt, Jianling Xie, Sarah A. Buchan, Jesse Papenburg, Jeffrey Cheng, Mohamed Eltorki, Yaron Finkelstein, Vikram Sabhaney, Jeffrey M. Pernica, Jocelyn Gravel, April J. Kam, Simon Berthelot, Brett Burstein, Jason Emsley, Ahmed Mater, Samina Ali, Archna Shah, Naveen Poonai, Darcy Beer, Peter J. Gill, James Dickinson, Stephen B. Freedman, the Surveillance Program for the Rapid IdeNtification, TracKing of Infectious Diseases in kidS (SPRINT-KIDS) Project Team

**Affiliations:** ICES, Public Health Ontario, Department of Family and Community Medicine and Dalla Lana School of Public Health, University of Toronto, Toronto, Ontario, Canada; Department of Pathology and Laboratory Medicine, BC Children’s Hospital, Vancouver, British Columbia, Canada; Division of Emergency Medicine, Children’s Hospital of Eastern Ontario, Ottawa, Ontario, Canada; Section of Pediatric Emergency Medicine, Department of Pediatrics, Cumming School of Medicine, University of Calgary, Calgary, AB, Canada; ICES, Public Health Ontario, Dalla Lana School of Public Health, University of Toronto, Toronto, Ontario, Canada; Division of Pediatric Infectious Diseases, Department of Pediatrics, Montreal Children’s Hospital, McGill University Health Centre, Montreal, Quebec, Canada; Section of Pediatric Emergency Medicine, Departments of Pediatrics and Emergency Medicine, Cumming School of Medicine, University of Calgary, Calgary, Alberta, Canada; Divisions of Paediatric Emergency Medicine and Clinical Pharmacology and Toxicology, The Hospital for Sick Children, University of Toronto, Toronto, Ontario, Canada; Division of Emergency Medicine, Department of Pediatrics, and BC Children’s Hospital Research Institute, Faculty of Medicine, University of British Columbia, Vancouver, British Columbia, Canada; Department of Pediatrics, Department of Health Research Methods, Evidence, and Impact, McMaster University, Hamilton, Ontario, Canada; Department of pediatric emergency medicine, Centre Hospitalier Universitaire Sainte-Justine, Université de Montréal, Montréal, Quebec, Canada; Division of Pediatric Emergency Medicine, Department of Pediatrics, McMaster University, Hamilton, Ontario, Canada; Département de médecine de famille et de médecine d’urgence, Faculté de médecine, Université Laval, Québec, Quebec, Canada; Montreal Children’s Hospital, Division of Pediatric Emergency Medicine, McGill University Health Centre, and Department of Epidemiology, Biostatistics and Occupational Health, McGill University, Montreal, Quebec, Canada; Department of Emergency Medicine, IWK Children’s Health Centre and QEII Health Sciences Centre, Dalhousie University, Halifax, Nova Scotia, Canada; Division of Pediatric Emergency, Department of Pediatrics, Jim Pattison Children’s Hospital, University of Saskatchewan, Saskatoon, Saskatchewan, Canada; Department of Pediatrics, Faculty of Medicine and Dentistry, University of Alberta, Edmonton, Alberta, Canada; Division of Pediatric Emergency Medicine, Discipline of Pediatrics, Memorial University, Janeway Children’s Health and Rehabilitation Center, NL Health Services, St Johns, NL, Canada; Departments of Paediatric and Epidemiology & Biostatistics, Schulich School of Medicine & Dentistry, Western University, London, Ontario, Canada; Department of Pediatrics and Child Health, Rady Faculty of Health Sciences, Max Rady College of Medicine, Winnipeg, MB, Canada; Department of Paediatrics, The Hospital for Sick Children, University of Toronto, Toronto, Ontario, Canada; Departments of Family Medicine and Community Health Sciences, University of Calgary, Calgary, Alberta, Canada; Sections of Pediatric Emergency Medicine and Gastroenterology, Departments of Pediatrics and Emergency Medicine, Cumming School of Medicine, University of Calgary, Calgary, Alberta, Canada

**Keywords:** Influenza, Immunization, Vaccine effectiveness, Children, Respiratory Tract Infections, Public Health

## Abstract

**Importance:** Influenza causes substantial morbidity in children, yet Canadian data on influenza vaccine effectiveness (VE) in preventing emergency department (ED) visits and hospitalizations are limited.

**Objective:** To estimate VE against influenza-associated ED visits and hospitalizations among children during the 2024-2025 and 2025-2026 seasons.

**Design:** Test-negative case-control design.

**Setting:** Fourteen Canadian pediatric tertiary-care hospitals participating in the Surveillance Program for the Rapid IdeNtification and TracKing of Infectious Diseases in kidS (SPRINT-KIDS) network.

**Participants:** Symptomatic children aged 6 months to <18 years tested for influenza from October 27, 2024, to June 7, 2025, and from November 2, 2025, to June 13, 2026.

**Exposure:** At least one dose of current-season influenza vaccine.

**Main Outcomes and Measures:** VE against laboratory-confirmed influenza A or B associated with an ED visit or hospitalization. VE was estimated using multivariable logistic regression adjusted for age, epidemiologic week, site, and high-risk comorbidity.

**Results:** The 2024-2025 season included 17,316 encounters (median [IQR] age=41.0 [18.5–82.3] months; 54.0% male), with 4,760 influenza-positive cases (8.9% vaccinated) and 12,556 influenza-negative controls (15.9% vaccinated). The 2025-2026 season included 13,417 encounters (median [IQR] age=33.5 [16.7–74.8] months; 54.1% male), with 4,798 cases (7.6% vaccinated) and 8,619 controls (18.1% vaccinated). VE against any influenza was 47% (95%CI, 40–53%) and 53% (95%CI, 46–59%) in 2024-2025 and 2025-2026, respectively. VE against ED visits was 53% (95%CI, 45–59%) and 57% (95%CI, 49–63%), and VE against hospitalization was 33% (95%CI, 16–47%) and 48% (95%CI, 33–59%) in 2024-2025 and 2025-2026, respectively. VE was lower against influenza A as compared to B in both seasons: 31% (95%CI, 22–39%) vs. 75% (95%CI, 64– 82%) (p<0.001) in 2024-2025; and 42% (95%CI, 32–50%) vs. 64% (95%CI, 55–72%) (p=0.001) in 2025-2026. VE was lower in children aged 10 to <18 years than in those <10 years, particularly among children with comorbidities. Protection against influenza A lasted up to 90 days, whereas it persisted beyond 120 days for influenza B.

**Conclusions and Relevance:** Influenza vaccines were moderately effective against pediatric ED visits and hospitalizations in both seasons. Routine vaccination should continue to be recommended to prevent hospital-attended influenza.

**Key Points:** *Question:* How effective were influenza vaccines against influenza-associated emergency department visits and hospitalizations among Canadian children during the 2024-2025 and 2025-2026 seasons?

*Findings:* In this test-negative case-control study of 30,733 encounters at 14 pediatric hospitals, vaccine effectiveness against any influenza was 47% in 2024-2025 and 53% in 2025-2026, despite antigenically drifted A(H3N2) subclade K predominance. Effectiveness was lower against influenza A than B and among adolescents.

*Meaning:* These findings support continued annual influenza vaccination of children, even when substantial antigenic mismatch is anticipated.

## INTRODUCTION

Influenza causes substantial morbidity in children. Hospitalization rates are highest among children aged <5 years, and up to 10% of pediatric hospitalizations during winter are attributable to influenza.^1,2^ Among Canadian children hospitalized with laboratory-confirmed influenza between 2012 and 2023, 17% required intensive care and 6% were mechanically ventilated.^3^ Emergency department (ED) visits substantially outnumber hospitalizations (e.g., 20-fold higher among children aged <5 years),^4^ yet remain less well-characterized. Since the resumption of typical influenza circulation after relaxation of COVID-19 pandemic restrictions, case numbers in children have approached pre-pandemic levels,^3,5^ while childhood vaccine coverage remains low (e.g., ranging between 30% and 34% during the 2021-2022 to 2023-2024 seasons in Canada).^6,7^

Annual vaccination is recommended for all children aged ³6 months, but vaccine effectiveness (VE) varies by season, influenza type and subtype, age, and product.^8,9^ The 2024-2025 Northern Hemisphere season featured co-circulation of A(H1N1)pdm09 and A(H3N2).^10^ The 2025-2026 season was dominated by A(H3N2) subclade K (J.2.4.1), which emerged after Northern Hemisphere vaccine strains were selected in February 2025.^11^ Epitope-based modeling and postvaccination neutralizing antibody studies suggested possibly reduced protection.^12–14^ Monitoring influenza VE is critical for guiding vaccination strategies and public health planning, and to inform expectations for the upcoming season.

Canadian VE estimates have historically been derived primarily from sentinel surveillance in physician offices.^15^ No surveillance platforms involving prospective data collection have estimated VE against influenza-associated pediatric ED visits or hospitalizations in Canada. The Surveillance Program for the Rapid IdeNtification and TracKing of Infectious Diseases in kidS (SPRINT-KIDS) network collects standardized clinical, virologic, and immunization data on children tested for respiratory viruses.^16^ Using SPRINT-KIDS data and a test-negative design, we estimated influenza VE against ED visits and hospitalizations among children aged 6 months to <18 years in Canada during the 2024-2025 and 2025-2026 influenza seasons, overall and by influenza type, disposition, age group, comorbidity status, time since vaccination, and vaccine product.

## METHODS

### Design and Setting

This multicenter test-negative case-control study used SPRINT-KIDS network data on children and adolescents tested for respiratory viruses at 14 pediatric tertiary-care hospitals in eight Canadian provinces (**eFigure 1**). Each site collected data with research ethics board (REB) approval and a waiver of consent or as an REB-exempt surveillance activity, in accordance with the Tri-Council Policy Statement: Ethical Conduct for Research Involving Humans.^17^ All analyses received REB approval. The study follows the STROBE reporting guideline (**eTable 1**).^18^

**Figure 1.**
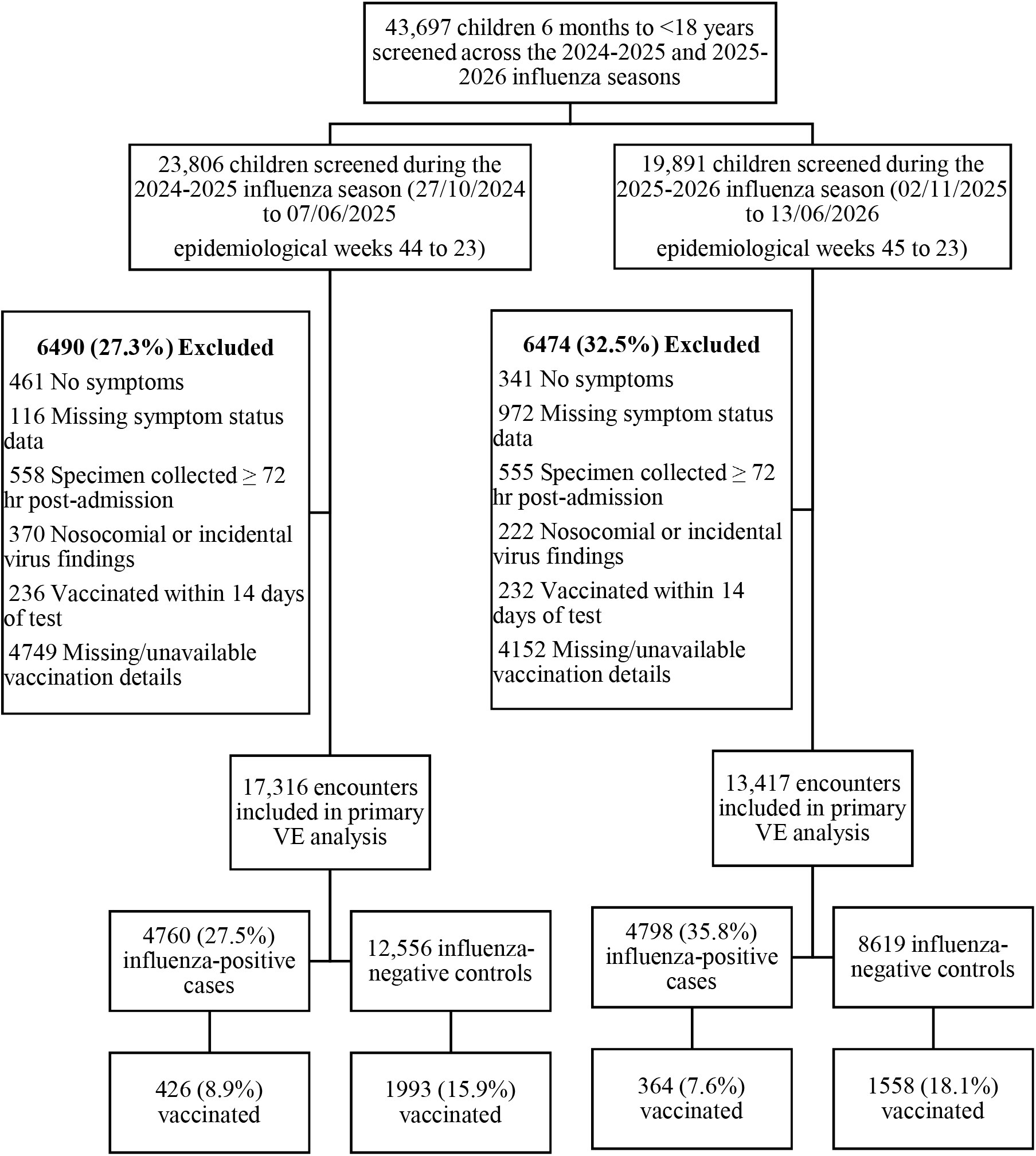
Flow diagram for study inclusion, restricting to children aged 6 months to <18 years tested for influenza, from October 27, 2024, to June 7, 2025 (epidemiological weeks 44 to 23) and from November 2, 2025, to June 13, 2026 (epidemiological weeks 45 to 23)

The surveillance program collects data on every child who tests positive for influenza, respiratory syncytial virus, and/or SARS-CoV-2 in the ED, inpatient units, or intensive care units (ICUs) of participating sites. To limit the burden of primary data collection, children who test negative for all three viruses (i.e., pan-negative controls) are randomly sampled each week at each site. The target is one pan-negative control for each case of the pathogen with the highest weekly case count at that site, so the controls represent eligible children with negative results. Treating clinicians decide which patients to test.

### Population

We included all children aged 6 months to <18 years who presented to a SPRINT-KIDS site who were tested for influenza by a nucleic acid amplification test (∼99%) or antigen test (∼1%) during influenza seasons, defined as consecutive weeks when >5% of influenza tests returned positive results, there were ≥15 cases in the surveillance network, and at least one tested child was considered vaccinated against influenza. The 2024-2025 season comprised epidemiologic weeks 44–23 (October 27, 2024, to June 7, 2025) and the 2025-2026 season weeks 45–23 (November 2, 2025, to June 13, 2026). We excluded children who were asymptomatic or had missing symptom data (to ensure detection reflected clinically relevant respiratory tract infections), were tested >72 hours after admission, had infections that were deemed nosocomial or incidental (i.e., unrelated to the reason for hospital admission), were vaccinated within 14 days of specimen collection, or had unknown influenza vaccination status. Children could contribute multiple episodes if subsequent testing occurred >14 days after a prior test.

### Exposure

Influenza vaccination was defined as receipt of ≥1 dose of influenza vaccine on or after October 1 of the corresponding season ≥14 days before specimen collection. Vaccination dates, number of current-season doses, and product type (i.e., inactivated or live-attenuated influenza vaccine [LAIV]) were identified from provincial immunization registries, medical records, or caregiver report, with registry or record documentation taking precedence when sources conflicted.

### Outcomes

The primary outcome was any laboratory-confirmed influenza A or B infection, irrespective of hospital setting. Secondary outcomes were defined by highest-level disposition within 14 days of the index test (i.e., ED visit only, non-ICU hospitalization, ICU admission). Each outcome was examined by influenza type (A or B), given their distinct antigenic and epidemiologic characteristics. Influenza A subtyping was performed at selected sites using clinically validated assays per local protocols.

### Covariates

We collected age, sex, province of residence, neighbourhood-level income derived from the forward sortation area (first 3 digits of the postal code), and high-risk comorbidities designated by Canada’s National Advisory Committee on Immunization as conferring increased risk for severe influenza, including cancer, immune-compromising condition or therapy, congenital heart disease, neurologic or neurodevelopmental conditions, chronic pulmonary disorders, and technology dependence.^9^

### Statistical analysis

For each season, we summarized baseline characteristics by test result and vaccination status using medians (IQR) for continuous and counts (%) for categorical variables. Differences between influenza-positive and influenza-negative groups, and between vaccinated and unvaccinated groups, were assessed using the Wilcoxon rank-sum test for continuous variables and the Chi-square test for categorical variables.

We used multivariable logistic regression to compare the odds of influenza vaccination in children with influenza-positive test results (i.e., cases) to those with influenza-negative test results (i.e., controls). VE was calculated as (1 − adjusted odds ratio [aOR]) × 100%. We adjusted for age group (6 to <24 months, 24 to <60 months, 5 to <10 years, 10 to <18 years), epidemiologic week, site, and presence of a high-risk comorbidity. Epidemiologic week was modeled using a natural cubic spline with 3 degrees of freedom to account for non-linear temporal trends. Analyses for the 2024-2025 and 2025-2026 seasons were conducted separately.

Because pan-negative controls were randomly sampled within site and calendar week independently of vaccination status, and children positive for other target viruses were enrolled completely, differential sampling fractions across control groups affect only control composition within each site-week stratum and not the within-stratum odds of vaccination. Adjustment for site and calendar week therefore accounts for the sampling design, and the aOR remains unbiased under the standard test-negative design assumption that influenza vaccination does not affect the risk of the other sampled viruses.^19^

We estimated VE against any influenza overall, and by disposition. For disposition-specific estimates, controls were restricted to test-negative children reaching the same level of care as the cases. Analyses were repeated by influenza type. Prespecified subgroup analyses estimated VE by age group, with further stratification by high-risk comorbidity. To assess waning of protection, we estimated VE by interval since most recent dose (14–30, 31–60, 61–90, 91–120, >120 days) separately for influenza A and B, because influenza B typically circulates later and interval since vaccination is otherwise confounded with calendar time. Among children aged ³2 years, the age at which LAIV is authorized in Canada, we estimated VE by influenza type for LAIV and inactivated vaccine. Further details are described in the **eMethods**.

We conducted sensitivity analyses to assess the impacts of 1) unavailable vaccination data; and 2) source of vaccination data. Details are provided in the **eMethods**.

Analyses used R 4.3.0 (R Foundation for Statistical Computing). All tests were 2-sided and used a significance level of *P*<0.05.

## RESULTS

Of 43,697 patient encounters screened among children aged 6 months to <18 years across the 2024-2025 and 2025-2026 influenza seasons, 30,733 were included. The most common reason for exclusion was missing vaccination data (n=8,901; 20.4%) (**Figure 1**). In both seasons, influenza A predominated and preceded influenza B, with greater separation of the A and B peaks in 2025-2026 (**Figure 2**). Subtyping information was available for 27.3% (1,009/3,700) of influenza A cases in 2024-2025 and 17.0% (603/3,560) in 2025-2026.

**Figure 2.**
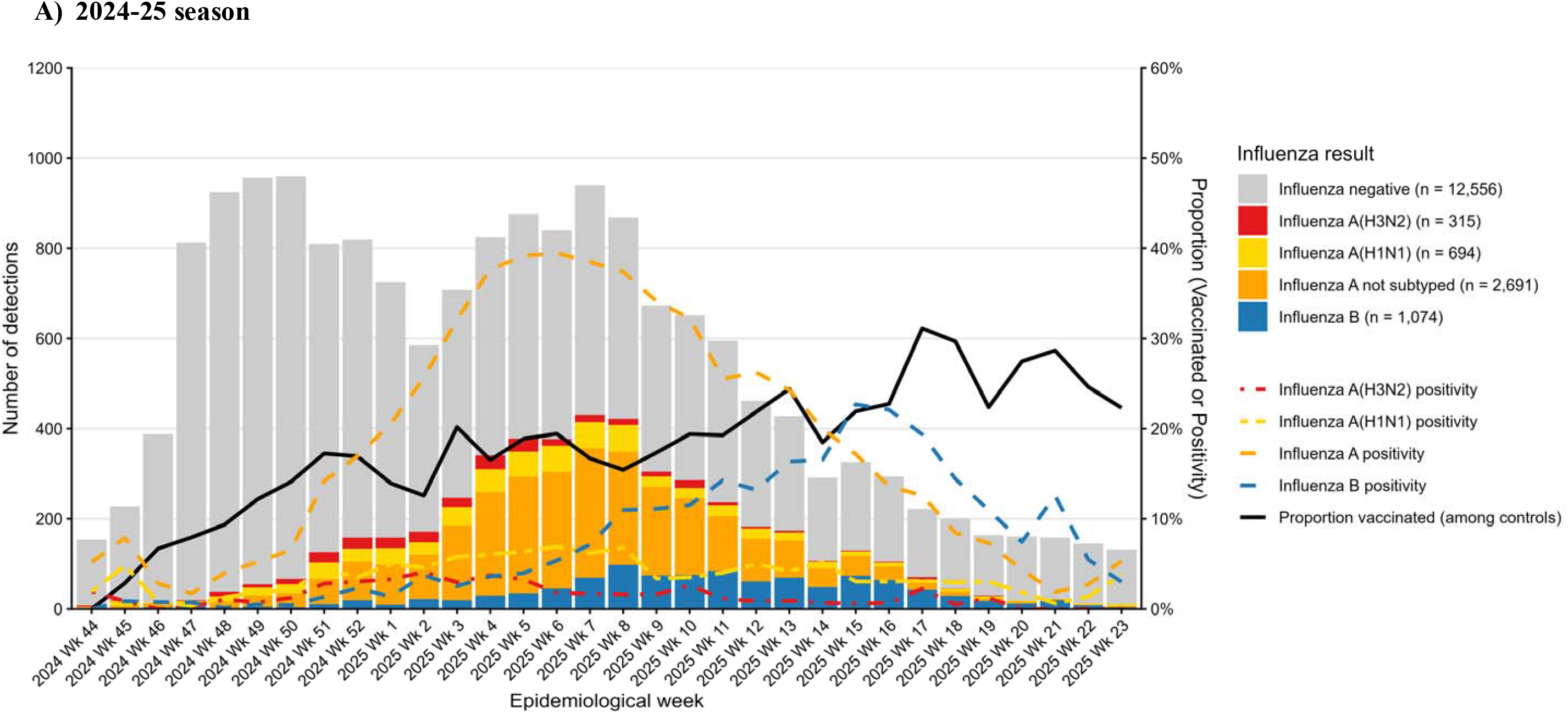
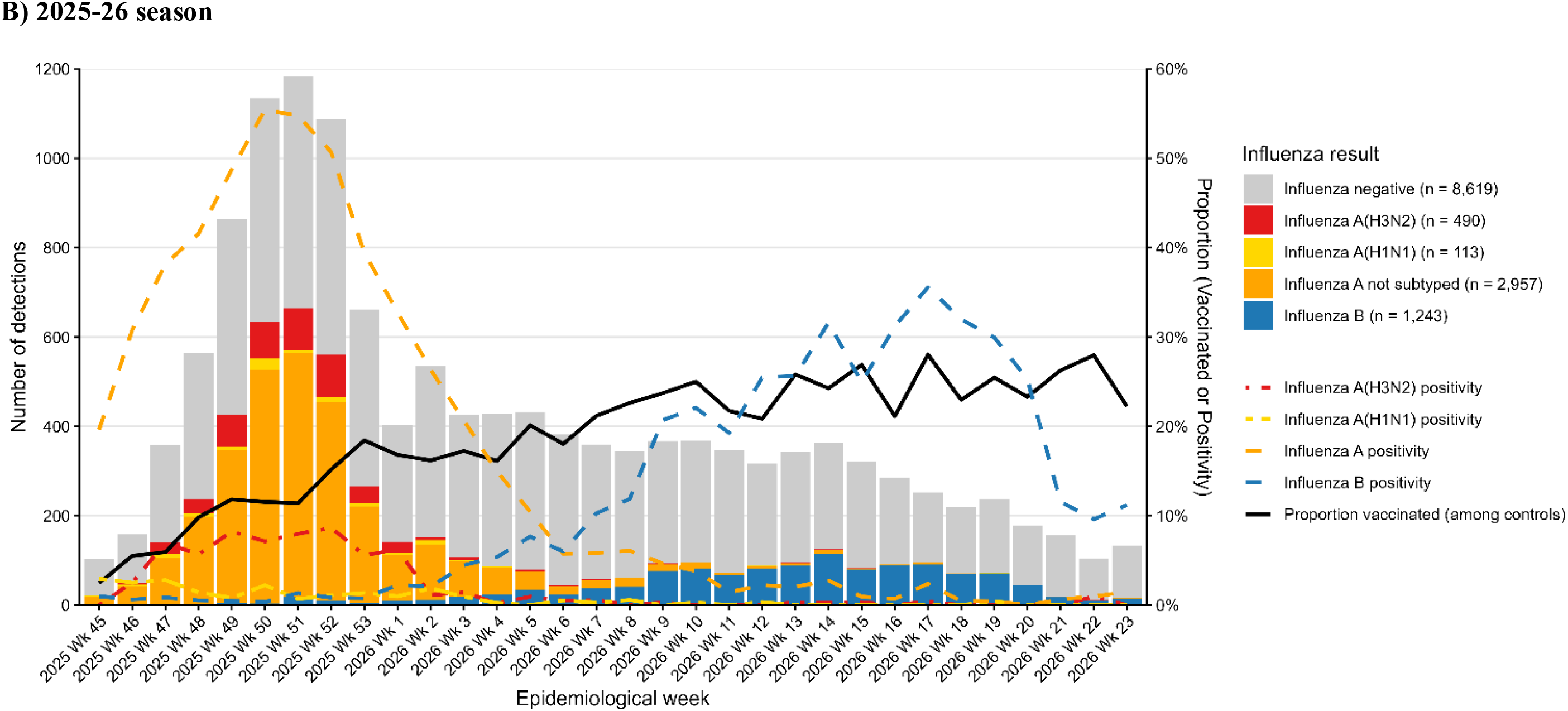
Influenza test-positive and test-negative specimens, positivity, and proportion of test-negative controls vaccinated, by week, from October 27, 2024, to June 7, 2025 (epidemiological weeks 44 to 23) (panel A) and from November 2, 2025, to June 13, 2026 (epidemiological weeks 45 to 22) (panel B)

Baseline characteristics are shown in **Table 1** (2024-2025) and **Table 2** (2025-2026). Overall, median age was 37.5 months (IQR 17.5–79.2) and 54.1% were male. In both seasons, compared with controls, cases were older, more likely to reside in British Columbia (BC), Ontario, or Quebec, less likely to have a comorbidity, more likely to be discharged from an ED, and less likely to be vaccinated. Among vaccinated children, cases had a longer interval since their last influenza vaccine dose than controls in 2024-2025 but a shorter interval in 2025-2026. In both seasons, compared with unvaccinated children, vaccinated children were younger, more likely to reside in higher-income neighbourhoods, more likely to be from Alberta or BC, and more likely to have a comorbidity.

**Table 1.** Characteristics of the study population by influenza test result and influenza vaccination status, 2024-2025 season.

| Variable, no. (%) | All<br>(N=17316) | Influenza-positive<br>(N=4760) | Influenza-negative<br>(N=12556) | P<br>value<sup>1</sup> | Vaccinated<br>(N=2419) | Unvaccinated<br>(N=14897) | P<br>value<sup>2</sup> |
|---|---|---|---|---|---|---|---|
| **Sociodemographic characteristics** |  |  |  |  |  |  |  |
| Age (months)<br>(median [IQR]) | 41.0<br>[18.5, 82.3] | 54.5<br>[27.8, 88.3] | 35.0<br>[16.5, 78.8] | <0.001 | 33.5<br>[16.2, 75.1] | 42.0<br>[18.9, 84.0] | <0.001 |
| Age group |  |  |  | <0.001 |  |  | <0.001 |
| 6 to <24 months | 5742 (33.2) | 969 (20.4) | 4773 (38.0) |  | 961 (39.7) | 4781 (32.1) |  |
| 24 to <60 months | 5228 (30.2) | 1652 (34.7) | 3576 (28.5) |  | 667 (27.6) | 4561 (30.6) |  |
| 5 to <10 years | 3866 (22.3) | 1511 (31.7) | 2355 (18.8) |  | 497 (20.5) | 3369 (22.6) |  |
| 10 to <18 years | 2480 (14.3) | 628 (13.2) | 1852 (14.7) |  | 294 (12.2) | 2186 (14.7) |  |
| Sex |  |  |  | 0.59 |  |  | 0.37 |
| Female | 7963 (46.0) | 2159 (45.4) | 5804 (46.2) |  | 1133 (46.8) | 6830 (45.8) |  |
| Male | 9350 (54.0) | 2600 (54.6) | 6750 (53.8) |  | 1285 (53.1) | 8065 (54.1) |  |
| Other | 3 (0.0) | 1 (0.0) | 2 (0.0) |  | 1 (0.0) | 2 (0.0) |  |
| Median total household income quintile (CAD) |  |  |  | 0.06 |  |  | <0.001 |
| \$40,800–71,500 | 3484 (20.2) | 1008 (21.3) | 2476 (19.8) |  | 356 (14.8) | 3128 (21.1) |  |
| \$71,501–83,000 | 3535 (20.5) | 968 (20.5) | 2567 (20.5) |  | 459 (19.0) | 3076 (20.8) |  |
| \$83,001–94,000 | 3435 (19.9) | 959 (20.3) | 2476 (19.8) |  | 509 (21.1) | 2926 (19.7) |  |
| \$94,001–110,000 | 3558 (20.6) | 968 (20.5) | 2590 (20.7) |  | 542 (22.5) | 3016 (20.3) |  |
| \$110,001–240,000 | 3222 (18.7) | 829 (17.5) | 2393 (19.1) |  | 547 (22.7) | 2675 (18.0) |  |
| Missing | 82 | 28 | 54 |  | 6 | 76 |  |
| Province of residence |  |  |  | <0.001 |  |  | <0.001 |
| Alberta | 4016 (23.2) | 645 (13.6) | 3371 (26.9) |  | 948 (39.2) | 3068 (20.6) |  |
| British Columbia | 2798 (16.2) | 980 (20.6) | 1818 (14.5) |  | 470 (19.4) | 2328 (15.6) |  |
| Manitoba | 653 (3.8) | 107 (2.2) | 546 (4.4) |  | 89 (3.7) | 564 (3.8) |  |
| NFLD & Labrador | 844 (4.9) | 152 (3.2) | 692 (5.5) |  | 80 (3.3) | 764 (5.1) |  |
| Nova Scotia | 611 (3.5) | 140 (2.9) | 471 (3.8) |  | 119 (4.9) | 492 (3.3) |  |
| Ontario | 3935 (22.7) | 1238 (26.0) | 2697 (21.5) |  | 363 (15.0) | 3572 (24.0) |  |
| Quebec | 4216 (24.4) | 1430 (30.0) | 2786 (22.2) |  | 335 (13.9) | 3881 (26.1) |  |
| Saskatchewan | 173 (1.0) | 55 (1.2) | 118 (0.9) |  | 6 (0.2) | 167 (1.1) |  |
| Others combined<sup>3</sup> | 63 (0.4) | 12 (0.3) | 51 (0.4) |  | 8 (0.3) | 55 (0.4) |  |
| Missing | 7 | 1 | 6 |  | 1 | 6 |  |
| **Clinical characteristics** |  |  |  |  |  |  |  |
| Diagnosed comorbidity<sup>4</sup> | 5766 (33.3) | 1416 (29.7) | 4350 (34.6) | <0.001 | 1039 (43.0) | 4727 (31.7) | <0.001 |
| Setting |  |  |  | <0.001 |  |  | 0.15 |
| ED visit only | 10165 (58.7) | 3565 (74.9) | 6600 (52.6) |  | 1377 (56.9) | 8788 (59.0) |  |
| Hospitalized<sup>5</sup> | 6173 (35.6) | 1017 (21.4) | 5156 (41.1) |  | 905 (37.4) | 5268 (35.4) |  |
| ICU | 978 (5.6) | 178 (3.7) | 800 (6.4) |  | 137 (5.7) | 841 (5.6) |  |
| **Influenza virus characteristics** |  |  |  |  |  |  |  |
| Influenza type |  |  |  |  |  |  |  |
| Influenza A | 3700 (21.4)<sup>6</sup> | 3700 (77.7) | 0 (0) | <0.001 | 389 (16.1) | 3311 (22.2) | <0.001 |
| Influenza B | 1074 (6.2)<sup>6</sup> | 1074 (22.6) | 0 (0) | <0.001 | 37 (1.5) | 1037 (7.0) | <0.001 |
| Month of test |  |  |  | <0.001 |  |  | <0.001 |
| October | 101 (0.6) | 5 (0.1) | 96 (0.8) |  | 0 (0.0) | 101 (0.7) |  |
| November | 2406 (13.9) | 95 (2.0) | 2311 (18.4) |  | 182 (7.5) | 2224 (14.9) |  |
| December | 3882 (22.4) | 481 (10.1) | 3401 (27.1) |  | 550 (22.7) | 3332 (22.4) |  |
| January | 3274 (18.9) | 1174 (24.7) | 2100 (16.7) |  | 450 (18.6) | 2824 (19.0) |  |
| February | 3320 (19.2) | 1532 (32.2) | 1788 (14.2) |  | 457 (18.9) | 2863 (19.2) |  |
| March | 2323 (13.4) | 945 (19.9) | 1378 (11.0) |  | 370 (15.3) | 1953 (13.1) |  |
| April | 1173 (6.8) | 416 (8.7) | 757 (6.0) |  | 219 (9.1) | 954 (6.4) |  |
| May | 705 (4.1) | 101 (2.1) | 604 (4.8) |  | 162 (6.7) | 543 (3.6) |  |
| June | 132 (0.8) | 11 (0.2) | 121 (1.0) |  | 29 (1.2) | 103 (0.7) |  |
| **Influenza vaccination characteristics** |  |  |  |  |  |  |  |
| Number of doses |  |  |  | <0.001 |  |  | <0.001 |
| Zero dose | 14897 (86.0) | 4334 (91.1) | 10563 (84.1) |  | 0 (0) | 14897 (100.0) |  |
| One dose | 2131 (12.3) | 404 (8.5) | 1727 (13.8) |  | 2131 (88.1) | 0 (0) |  |
| Two doses | 288 (1.7) | 22 (0.5) | 266 (2.1) |  | 288 (11.9) | 0 (0) |  |
| Type of vaccine |  |  |  | 0.002 |  |  |  |
| Inactivated | 1952 (80.7) | 325 (76.3) | 1627 (81.6) |  | 1952 (80.7) | N/A |  |
| Live-attenuated | 98 (4.1) | 30 (7.0) | 68 (3.4) |  | 98 (4.1) | N/A |  |
| Unknown | 369 (15.3) | 71 (16.7) | 298 (15.0) |  | 369 (15.3) | N/A |  |
| Days since last dose<br>(median [IQR]) | 66.0 [39.0, 103.0] | 87.0 [56.0, 109.0] | 62.0 [37.0, 101.0] | <0.001 | 66.0 [39.0, 103.0] | N/A |  |

<sup>1</sup>Comparing influenza test result. <sup>2</sup>Comparing influenza vaccination status. <sup>3</sup>Yukon, Northwest Territories, Nunavut, New Brunswick, Prince Edward Island, and non-Canadian residents. <sup>4</sup>Comorbidity was defined as any presence of: cancer/immune compromising condition, congenital heart disease, neurologic or neurodevelopment conditions, pulmonary disorders, and technology dependence. <sup>5</sup>Does not include admission to intensive care. <sup>6</sup>Includes 14 influenza A and B co-infections.
Abbreviations: ED = emergency department; ICU = intensive care unit; IQR = interquartile range; NFLD = Newfoundland.

**Table 2.** Characteristics of the study population by influenza test result and influenza vaccination status, 2025-2026 season.

| Variable, no. (%) | All<br>(N=13417) | Influenza-positive<br>(N=4798) | Influenza-negative<br>(N=8619) | P value<sup>1</sup> | Vaccinated<br>(N=1922) | Unvaccinated<br>(N=11495) | P value<sup>2</sup> |
|---|---|---|---|---|---|---|---|
| **Sociodemographic characteristics** |  |  |  |  |  |  |  |
| Age (months) (median [IQR]) | 33.5 [16.7, 74.8] | 51.4 [24.5, 93.4] | 26.3 [14.7, 58.7] | <0.001 | 27.4 [15.4, 74.8] | 34.5 [17.0, 74.8] | <0.001 |
| Age group |  |  |  | <0.001 |  |  | <0.001 |
| 6 to <24 months | 5197 (38.7) | 1177 (24.5) | 4020 (46.6) |  | 872 (45.4) | 4325 (37.6) |  |
| 24 to <60 months | 4063 (30.3) | 1565 (32.6) | 2498 (29.0) |  | 471 (24.5) | 3592 (31.2) |  |
| 5 to <10 years | 2318 (17.3) | 1253 (26.1) | 1065 (12.4) |  | 338 (17.6) | 1980 (17.2) |  |
| 10 to <18 years | 1839 (13.7) | 803 (16.7) | 1036 (12.0) |  | 241 (12.5) | 1598 (13.9) |  |
| Sex |  |  |  | 0.64 |  |  | 0.65 |
| Female | 6158 (45.9) | 2189 (45.6) | 3969 (46.0) |  | 892 (46.4) | 5267 (45.8) |  |
| Male | 7258 (54.1) | 2609 (54.4) | 4649 (53.9) |  | 1030 (53.6) | 6228 (54.2) |  |
| Missing | 1 | 0 | 1 |  | 0 | 1 |  |
| Median total household income quintile (CAD) |  |  |  | 0.001 |  |  | <0.001 |
| \$36,400–71,000 | 2730 (20.4) | 1065 (22.3) | 1665 (19.4) |  | 266 (13.9) | 2464 (21.5) |  |
| \$71,001–83,000 | 2628 (19.7) | 914 (19.1) | 1714 (20.0) |  | 328 (17.1) | 2300 (20.1) |  |
| \$83,001–96,000 | 2953 (22.1) | 1007 (21.1) | 1946 (22.7) |  | 505 (26.3) | 2448 (21.4) |  |
| \$96,001–110,000 | 2433 (18.2) | 845 (17.7) | 1588 (18.5) |  | 355 (18.5) | 2078 (18.2) |  |
| \$110,001–240,000 | 2623 (19.6) | 951 (19.9) | 1672 (19.5) |  | 464 (24.2) | 2159 (18.9) |  |
| Missing | 50 | 16 | 34 |  | 4 | 46 |  |
| Province/territory of residence |  |  |  | <0.001 |  |  | <0.001 |
| Alberta | 3625 (27.0) | 776 (16.2) | 2849 (33.1) |  | 858 (44.6) | 2767 (24.1) |  |
| British Columbia | 917 (6.8) | 378 (7.9) | 539 (6.3) |  | 169 (8.8) | 748 (6.5) |  |
| Manitoba | 505 (3.8) | 132 (2.8) | 373 (4.3) |  | 71 (3.7) | 434 (3.8) |  |
| NFLD & Labrador | 773 (5.8) | 273 (5.7) | 500 (5.8) |  | 149 (7.8) | 624 (5.4) |  |
| Nova Scotia | 537 (4.0) | 270 (5.6) | 267 (3.1) |  | 106 (5.5) | 431 (3.8) |  |
| Ontario | 3292 (24.5) | 1444 (30.1) | 1848 (21.4) |  | 270 (14.0) | 3022 (26.3) |  |
| Quebec | 3403 (25.4) | 1380 (28.8) | 2023 (23.5) |  | 293 (15.2) | 3110 (27.1) |  |
| Saskatchewan | 291 (2.2) | 122 (2.5) | 169 (2.0) |  | 4 (0.2) | 287 (2.5) |  |
| Others combined<sup>1</sup> | 72 (0.5) | 22 (0.5) | 50 (0.6) |  | 2 (0.1) | 70 (0.6) |  |
| Missing | 2 | 1 | 1 |  | 1 | 1 |  |
| **Clinical characteristics** |  |  |  |  |  |  |  |
| Diagnosed comorbidity<sup>2</sup> | 4554 (33.9) | 1508 (31.4) | 3046 (35.3) | <0.001 | 826 (43.0) | 3728 (32.4) | <0.001 |
| Setting |  |  |  | <0.001 |  |  | 0.02 |
| ED visit only | 7691 (57.3) | 3502 (73.0) | 4189 (48.6) |  | 1046 (54.4) | 6645 (57.8) |  |
| Hospitalized (not ICU) | 4825 (36.0) | 1085 (22.6) | 3740 (43.4) |  | 735 (38.2) | 4090 (35.6) |  |
| ICU | 901 (6.7) | 211 (4.4) | 690 (8.0) |  | 141 (7.3) | 760 (6.6) |  |
| **Influenza virus characteristics** |  |  |  |  |  |  |  |
| Influenza A | 3560 (26.5)<sup>6</sup> | 3560 (74.2) | 0 (0) | <0.001 | 265 (13.8) | 3295 (28.7) | <0.001 |
| Influenza B | 1243 (9.3)<sup>6</sup> | 1243 (25.9) | 0 (0) | <0.001 | 101 (5.3) | 1142 (9.9) | <0.001 |
| Month of test |  |  |  | <0.001 |  |  | <0.001 |
| November | 1299 (9.7) | 502 (10.5) | 797 (9.2) |  | 72 (3.7) | 1227 (10.7) |  |
| December | 4566 (34.0) | 2410 (50.2) | 2156 (25.0) |  | 458 (23.8) | 4108 (35.7) |  |
| January | 2039 (15.2) | 570 (11.9) | 1469 (17.0) |  | 303 (15.8) | 1736 (15.1) |  |
| February | 1519 (11.3) | 243 (5.1) | 1276 (14.8) |  | 283 (14.7) | 1236 (10.8) |  |
| March | 1552 (11.6) | 385 (8.0) | 1167 (13.5) |  | 301 (15.7) | 1251 (10.9) |  |
| April | 1344 (10.0) | 424 (8.8) | 920 (10.7) |  | 276 (14.4) | 1068 (9.3) |  |
| May | 881 (6.6) | 236 (4.9) | 645 (7.5) |  | 178 (9.3) | 703 (6.1) |  |
| June | 217 (1.6) | 28 (0.6) | 189 (2.2) |  | 51 (2.7) | 166 (1.4) |  |
| **Influenza vaccination characteristics** |  |  |  |  |  |  |  |
| Number of doses |  |  |  | <0.001 |  |  | <0.001 |
| Zero dose | 11495 (85.7) | 4434 (92.4) | 7061 (81.9) |  | 0 (0) | 11495 (100.0) |  |
| One dose | 1637 (12.2) | 345 (7.2) | 1292 (15.0) |  | 1637 (85.2) | 0 (0) |  |
| Two doses | 285 (2.1) | 19 (0.4) | 266 (3.1) |  | 285 (14.8) | 0 (0) |  |
| Type of vaccine |  |  |  | <0.001 |  |  |  |
| Inactivated | 1635 (85.1) | 287 (78.8) | 1348 (86.5) |  | 1635 (85.1) | N/A |  |
| Live-attenuated | 39 (2.0) | 8 (2.2) | 31 (2.0) |  | 39 (2.0) | N/A |  |
| Unknown | 248 (12.9) | 69 (19.0) | 179 (11.5) |  | 248 (12.9) | N/A |  |
| Days since last dose (median [IQR]) | 69.0 [39.0, 113.8] | 55.0 [36.0, 97.0] | 71.5 [40.0, 116.0] | <0.001 | 69.0 [39.0, 113.8] | N/A |  |

<sup>1</sup>Comparing influenza test result. <sup>2</sup>Comparing influenza vaccination status. <sup>3</sup>Yukon, Northwest Territories, Nunavut, New Brunswick, Prince Edward Island, and non-Canadian residents. <sup>4</sup>Comorbidity was defined as any presence of: cancer/immune compromising condition, congenital heart disease, neurologic or neurodevelopment conditions, pulmonary disorders, and technology dependence. <sup>6</sup>Includes 5 influenza A and B co-infections.
Abbreviations: ED = emergency department; ICU = intensive care unit; IQR = interquartile range; NFLD = Newfoundland.

VE against any influenza was similar across seasons: 47% (95%CI, 40–53%) in 2024-2025 and 53% (95%CI, 46–59%) in 2025-2026 (**Figure 3a**). VE was 53% (95%CI, 45–59%) and 57% (95%CI, 49–63%) against ED visits, and 33% (95%CI, 16–47%) and 48% (95%CI, 33–59%) against hospitalizations. VE was lower against influenza A than B: 31% (95%CI, 22–39%) vs. 75% (95%CI, 64–82%) (p<0.001) in 2024-2025 and 42% (95%CI, 32–50%) vs. 64% (95%CI, 55–72%) (p=0.001) in 2025-2026. (**Figures 3b** and **3c**).

**Figure 3.**
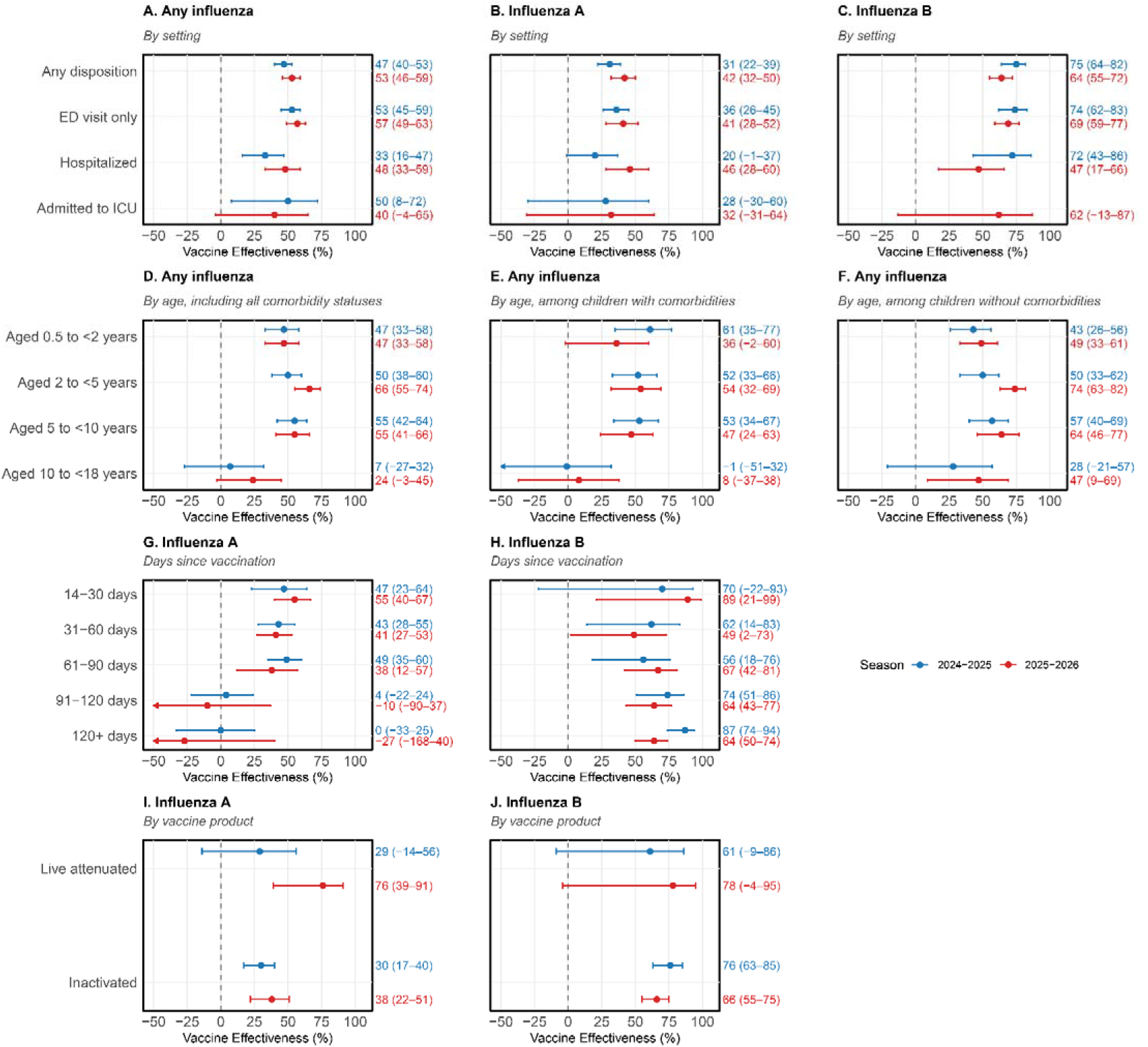
Estimates of influenza vaccine effectiveness against any influenza (A or B) in any setting and by disposition (panel A), against influenza A by disposition (panel B), against influenza B by disposition (panel C), against any influenza by age (panel D), against any influenza by age in children with comorbidities (panel E), against any influenza by age in children without comorbidities (panel F), against influenza A by time since vaccination (panel G), against influenza B by time since vaccination (panel H), against influenza A by vaccine product (panel I), against influenza B by vaccine product (panel J), for the 2024-2025 (red) and 2025-2026 (blue) seasons

VE was lower for children aged 10 to <18 years than for those aged 6 months to <10 years (percentage point difference=45 [95%CI 15–75]) in 2024-2025 and 31 [95%CI 10–52] in 2025-2026; **Figure 3d**). This pattern was more pronounced among children with (34 to 66 percentage point difference; **Figure 3e**) than without (16 to 17 percentage point difference; **Figure 3f**) a high-risk comorbidity.

VE against influenza A was consistent (43–49%) within 90 days of vaccination in 2024-2025 and declined from 55% to 38% in 2025-2026 (**Figure 3g**). Beyond 90 days, point estimates were lower, with wider 95%CIs that crossed the null in both seasons. VE against influenza B remained elevated beyond 120 days (**Figure 3h**).

LAIV and inactivated vaccine provided similar protection against influenza A in 2024-2025, but in 2025-2026, VE was higher for LAIV (76%; 95%CI, 43–94%) than for inactivated vaccine (38%; 95%CI, 23–51%), a difference of 38 percentage points (95%CI, 2–61) (p=0.04) (**Figure 3i**). No differences in VE between LAIV and inactivated vaccine were observed against influenza B in either season (**Figure 3j**).

The province-specific proportions of children with unavailable vaccination status are presented in **eTable 2**. Characteristics of children excluded for missing vaccination status are presented in **eTables 3** and **4**. Vaccination status was derived mainly from vaccine registries in all provinces except Ontario and Saskatchewan (**eTable 5**). The sensitivity analyses were consistent with the primary analysis (**eTable 6**).

## DISCUSSION

In this multicenter test-negative study of over 30,000 encounters across 14 Canadian pediatric tertiary care hospitals, influenza vaccination was associated with moderate protection against hospital-attended influenza during the 2024-2025 and 2025-2026 seasons, including when the antigenically drifted A(H3N2) subclade K (J.2.4.1) predominated in the latter season. Four findings warrant emphasis: 1) VE was consistently lower against influenza A than B; 2) VE was lower among children aged 10 to <18 years, particularly those with high-risk comorbidities; 3) protection against influenza A appeared lower beyond 90 days, whereas protection against influenza B persisted beyond 120 days post vaccination; and 4) in 2025-2026, VE against influenza A was higher for LAIV than inactivated vaccine.

Our estimates of VE against influenza A for 2025-2026 are consistent with international pediatric evidence that subclade K drift did not translate into reduced clinical protection, with our estimate against ED visits of 41% (95%CI, 28–52%) quite similar to estimates from the US against ED and urgent care encounters at 37% (95%CI, 32–41%)^12^ and from Canada against primary care visits at 35% (95%CI, 10–54%).^15^ Our results are also comparable to, or somewhat lower than, VE estimates from Europe against primary care and ED visits, which ranged from 47% (95%CI, 20– 76%) to 60% (95%CI, 51–66%).^20^ Our estimate of VE against hospitalization of 46% (95%CI, 28– 60%) is similar to an estimate from the US at 47% (95%CI, 14–69%)^12^ but lower than estimates from Europe, which ranged from 61% (95%CI, 41–74%) to 72% (95%CI, 67–77%).^20^ Human serologic studies suggest that vaccination and pre-existing immunity may provide clinical protection against subclade K despite the degree of antigenic drift suggested by laboratory models.^12^ ^21^

Pediatric VE against A(H3N2) has consistently been shown to be lower than against A(H1N1)pdm09 and influenza B, but VE against influenza B may be higher or lower than against A(H1N1)pdm09 depending on the season,^22–26^ likely reflecting differences in circulating and vaccine strains. The lower VE point estimates among adolescents compared to younger children, particularly those with high-risk comorbidities, is consistent with data from certain prior seasons^22–24^ but not other seasons.^24,27^ At least two non-mutually-exclusive mechanisms may contribute. First, immune imprinting and cumulative exposure differ by age, and antibody landscapes are shaped by early-life priming, such that responses to updated A(H3N2) vaccine antigens may be redirected toward previously encountered strains.^28^ Second, older children more often received multiple prior vaccinations, and repeat annual vaccination has been associated with attenuated VE, particularly against A(H3N2).^25^ Further research to elucidate the mechanisms for these findings is warranted.

Our finding of lower influenza A VE point estimates beyond 90 days should be interpreted cautiously against largely reassuring pediatric literature.^29^ A five-season US New Vaccine Surveillance Network analysis found only minimal, nonsignificant within-season decline in VE against hospitalization, concluding vaccination on current ACIP timing remains optimal.^26^ The American Academy of Pediatrics notes observed waning has been inconsistent across age groups and is more evident against A(H3N2).^7^ Several features argue against over-interpretation. Although we estimated VE separately by type, and genetic drift of influenza A viruses over the course of a season may be possible,^30,31^ residual confounding by calendar time and systematic differences between early and late vaccinees cannot be excluded. In test-negative waning analyses, depletion of susceptibles and partial vaccine protection can each generate apparent declines even when true per-exposure protection is stable.^32^ We therefore regard the waning influenza A VE signal as hypothesis-generating.

The higher VE of the LAIV against influenza A as compared to the inactivated vaccine during the A(H3N2)-predominant 2025-2026 season, but not the A(H1N1)pdm09-predominant 2024-2025 season, is biologically plausible. It is concordant with early-season 2025-2026 data from England, where children aged 2–17 years primarily receive LAIV, and pediatric VE against influenza A hospitalization was 74% (95%CI, 63–82%).^33^ Similarly, VE against influenza A was 81% (95%CI, 15–96%) for LAIV and 43% (95%CI, 15–62%) for inactivated vaccine during the 2025-2026 season in Japan.^34^ Previous differences in the opposite direction seen in the US in 2013-2014 through 2015-2016, with markedly lower LAIV4 effectiveness (20%) than inactivated vaccine (67%) for A(H1N1)pdm09,^35^ may have been related to reduced replicative fitness and increased thermolability of the A(H1N1)pdm09 LAIV vaccine strain that was subsequently changed.^36^ In contrast, Canadian data from 2012-2013 through 2015-2016 found no significant overall difference between products in children, including against A(H3N2).^37^ Given the small number of LAIV recipients, wide confidence intervals, and absence of subtype-level data, our product comparison is hypothesis-generating and not a basis for programmatic change.

The large sample size, multi-site surveillance design, and assessment of VE across two consecutive influenza seasons strengthen the robustness and relevance of the findings. Nevertheless, this study has some limitations. First, 20% of screened patients were excluded due to unavailable vaccination data. Reassuringly, our sensitivity analysis excluding data from jurisdictions with high proportions of children missing vaccination data demonstrated similar findings. Second, vaccination status was ascertained from registries, medical records, or caregiver report, with reliance on caregiver report varying by site. Our sensitivity analyses restricting the exposure definition to registry- or medical record-documented vaccination confirmed that our primary analysis was robust. Furthermore, Canadian sentinel physician VE estimates based on self-reported vaccination status versus a provincial registry were nearly identical.^38^ Third, influenza A subtyping was performed only at selected sites and was available for a minority of influenza A cases, preventing estimation of subtype-specific VE. Because subtyping was not random, reported subtype proportions may not be representative, and our conclusions regarding protection during subclade K circulation are ecological rather than VE against subclade K per se. Within-subtype heterogeneity could be masked by pooling, with uncertain direction of bias. Fourth, test-negative controls were sampled rather than fully enumerated. Unmeasured differences between sampled and unsampled controls cannot be excluded. Although disposition-matching aligned cases and controls on level of care, residual confounding by unmeasured determinants of admission may persist. Fifth, as an observational study, residual confounding by health-promoting behaviors, underlying health status, and unmeasured socioeconomic factors remains possible. Sixth, children could contribute more than one episode, and because encounters were not linked individually, we could not account for within-child correlation. As this influences precision rather than central tendency, we do not expect a materially important effect on our conclusions. Finally, these estimates derive from tertiary pediatric centers and may not generalize to community hospitals or mixed EDs.

In conclusion, among children presenting to Canadian pediatric tertiary care hospitals, influenza vaccination was associated with ∼50% lower odds of hospital-attended influenza during the 2024-2025 and 2025-2026 seasons, including during a season dominated by the antigenically drifted A(H3N2) subclade K. Lower VE among older children, particularly those with comorbidities, and against influenza A compared with influenza B, highlights the importance of continued surveillance and targeted prevention efforts. These findings support continued annual influenza vaccination of children even when substantial antigenic mismatch is anticipated.

## Data Availability

Data dictionary and analytic code will be shared, upon reasonable request, for academic purposes, with appropriate individuals who have obtained appropriate ethics permissions and data sharing agreements. Due to privacy requirements data cannot be shared.

## Acknowledgements

We thank our colleagues at the Public Health Agency of Canada for their collaboration on this project and all the SPRINT-KIDS project team members at our sites for their assistance.

## Additional Contributions

We thank our colleagues at the Public Health Agency of Canada for their collaboration on this project and all the research assistants, coordinators and nurses who contributed to data collection at all participating sites. We also thank the analysts at Alberta’s Public Health Surveillance & Informatics, Health Shared Services.

