## Supplemental Appendix for "Pediatric Influenza Vaccine Effectiveness Against Emergency Visits and Hospitalization in Canada: 2024-2025 and 2025-2026 seasons"

**Online-Only Supplement**

**eMethods.**	Statistical analysis.

**eTable 1.**	STROBE reporting guideline.

**eTable 2.**	Proportion of children with unavailable vaccination data.

### **eTable 3**. Comparison of baseline characteristics between included and those excluded due to missing vaccination status, 2024-2025 season.

### **eTable 4**. Comparison of baseline characteristics between included children and those excluded due to missing vaccination status, 2025-2026 season.

**eTable 5.**	Proportions of vaccinated children with vaccination status determined by provincial immunization registries, medical records, and caregiver report, by influenza season and province.

**eTable 6.**	Sensitivity analyses of vaccine effectiveness (VE) against any influenza in any setting for the 2024-2025 and 2025-2026 seasons.

**eMethods**

**Statistical Analysis**

Vaccine effectiveness by influenza type

To compare vaccine effectiveness (VE) by influenza type, separate adjusted logistic regression models were fitted using the same set of testing encounters, with influenza A and B positivity as the respective outcomes. We used a bootstrap procedure with 2000 resamples of testing encounters to estimate the uncertainty in the difference in VE. Both models were refit to the same bootstrap sample in each replicate to preserve dependence between estimates. For each replicate, VE was calculated for each influenza type, and the difference in VE (influenza A minus influenza B) was calculated. A 95% confidence interval (CI) for this difference was obtained from the bootstrap distribution. A two-sided P value was calculated using a normal approximation based on the observed difference in the vaccination coefficients on the log-odds scale and its bootstrap standard error.

Vaccine effectiveness by vaccine product

In the analysis comparing VE for LAIV and inactivated vaccine, we used three exposure groups (unvaccinated, LAIV, inactivated vaccine).The difference in VE was calculated as the VE for LAIV minus the VE for inactivated vaccine. Confidence intervals for VE and the VE difference were estimated using a participant-level nonparametric bootstrap with 2000 resamples drawn with replacement. The regression model was refitted in each resample, and 95% CIs were calculated using the 2.5^th^ and 97.5^th^ percentiles of the bootstrap estimates. Differences between vaccine type effects were assessed using a likelihood-ratio test comparing the model with separate coefficients for LAIV and inactivated vaccine with a reduced model imposing a common coefficient for both vaccinated groups.

Sensitivity analyses

First, to assess the impact of missing vaccination status, we excluded children from provinces and territories with high levels of unavailable vaccination data, defined initially as >20% of children with unavailable data, and subsequently as >10% of children with unavailable data.

Second, because reliance on caregiver report versus documentation varied by site, we restricted the exposure definition to registry- or medical record-documented vaccination (i.e., re-classifying caregiver-report-only as unvaccinated, and separately excluding them) to assess the impact of differential ascertainment.

**eTable 1. STROBE reporting guideline**

STROBE Statement—Checklist of items that should be included in reports of *case-control studies*

|  | Item No | Recommendation | Page No |
| --- | --- | --- | --- |
| Title and abstract | 1 | (*a*) Indicate the study’s design with a commonly used term in the title or the abstract | 6 |
|  |  | (*b*) Provide in the abstract an informative and balanced summary of what was done and what was found | 6 |
| Introduction | | | |
| Background/rationale | 2 | Explain the scientific background and rationale for the investigation being reported | 8 |
| Objectives | 3 | State specific objectives, including any prespecified hypotheses | 8-9 |
| Methods | | | |
| Study design | 4 | Present key elements of study design early in the paper | 9 |
| Setting | 5 | Describe the setting, locations, and relevant dates, including periods of recruitment, exposure, follow-up, and data collection | 9 |
| Participants | 6 | (*a*) Give the eligibility criteria, and the sources and methods of case ascertainment and control selection. Give the rationale for the choice of cases and controls | 9-10, 11 |
|  |  | (*b*) For matched studies, give matching criteria and the number of controls per case | N/A |
| Variables | 7 | Clearly define all outcomes, exposures, predictors, potential confounders, and effect modifiers. Give diagnostic criteria, if applicable | 10-11 |
| Data sources/ measurement | 8* | For each variable of interest, give sources of data and details of methods of assessment (measurement). Describe comparability of assessment methods if there is more than one group | 10-11 |
| Bias | 9 | Describe any efforts to address potential sources of bias | 11-12, eMethods |
| Study size | 10 | Explain how the study size was arrived at | N/A |
| Quantitative variables | 11 | Explain how quantitative variables were handled in the analyses. If applicable, describe which groupings were chosen and why | 11-12 |
| Statistical methods | 12 | (*a*) Describe all statistical methods, including those used to control for confounding | 11-12, eMethods |
|  |  | (*b*) Describe any methods used to examine subgroups and interactions | 12 |
|  |  | (*c*) Explain how missing data were addressed | 11-12, eMethods |
|  |  | (*d*) If applicable, explain how matching of cases and controls was addressed | N/A |
|  |  | (*e*) Describe any sensitivity analyses | 12,  eMethods |
| Results | | | |
| Participants | 13* | (a) Report numbers of individuals at each stage of study—eg numbers potentially eligible, examined for eligibility, confirmed eligible, included in the study, completing follow-up, and analysed | 12,  Figure 1 |
|  |  | (b) Give reasons for non-participation at each stage | Figure 1 |
|  |  | (c) Consider use of a flow diagram | Figure 1 |
| Descriptive data | 14* | (a) Give characteristics of study participants (eg demographic, clinical, social) and information on exposures and potential confounders | 13,Tables 1 & 2 |
|  |  | (b) Indicate number of participants with missing data for each variable of interest | Tables 1 & 2, eTable 2 |
| Outcome data | 15* | Report numbers in each exposure category, or summary measures of exposure | Tables 1 & 2 |

| Main results | 16 | (*a*) Give unadjusted estimates and, if applicable, confounder-adjusted estimates and their precision (eg, 95% confidence interval). Make clear which confounders were adjusted for and why they were included | 13, Figure 3 |
| --- | --- | --- | --- |
|  |  | (*b*) Report category boundaries when continuous variables were categorized | Tables 1 & 2, Figure 3 |
|  |  | (*c*) If relevant, consider translating estimates of relative risk into absolute risk for a meaningful time period | N/A |
| Other analyses | 17 | Report other analyses done—eg analyses of subgroups and interactions, and sensitivity analyses | 13-14,  eTable 6 |
| Discussion | | | |
| Key results | 18 | Summarise key results with reference to study objectives | 14 |
| Limitations | 19 | Discuss limitations of the study, taking into account sources of potential bias or imprecision. Discuss both direction and magnitude of any potential bias | 16-17 |
| Interpretation | 20 | Give a cautious overall interpretation of results considering objectives, limitations, multiplicity of analyses, results from similar studies, and other relevant evidence | 14-18 |
| Generalisability | 21 | Discuss the generalisability (external validity) of the study results | 17 |
| Other information | | | |
| Funding | 22 | Give the source of funding and the role of the funders for the present study and, if applicable, for the original study on which the present article is based | 4 |

*Give information separately for cases and controls.

Note: An Explanation and Elaboration article discusses each checklist item and gives methodological background and published examples of transparent reporting. The STROBE checklist is best used in conjunction with this article (freely available on the Web sites of PLoS Medicine at http://www.plosmedicine.org/, Annals of Internal Medicine at http://www.annals.org/, and Epidemiology at http://www.epidem.com/). Information on the STROBE Initiative is available at http://www.strobe-statement.org.

**eFigure 1. Map of SPRINT-KIDS sites**

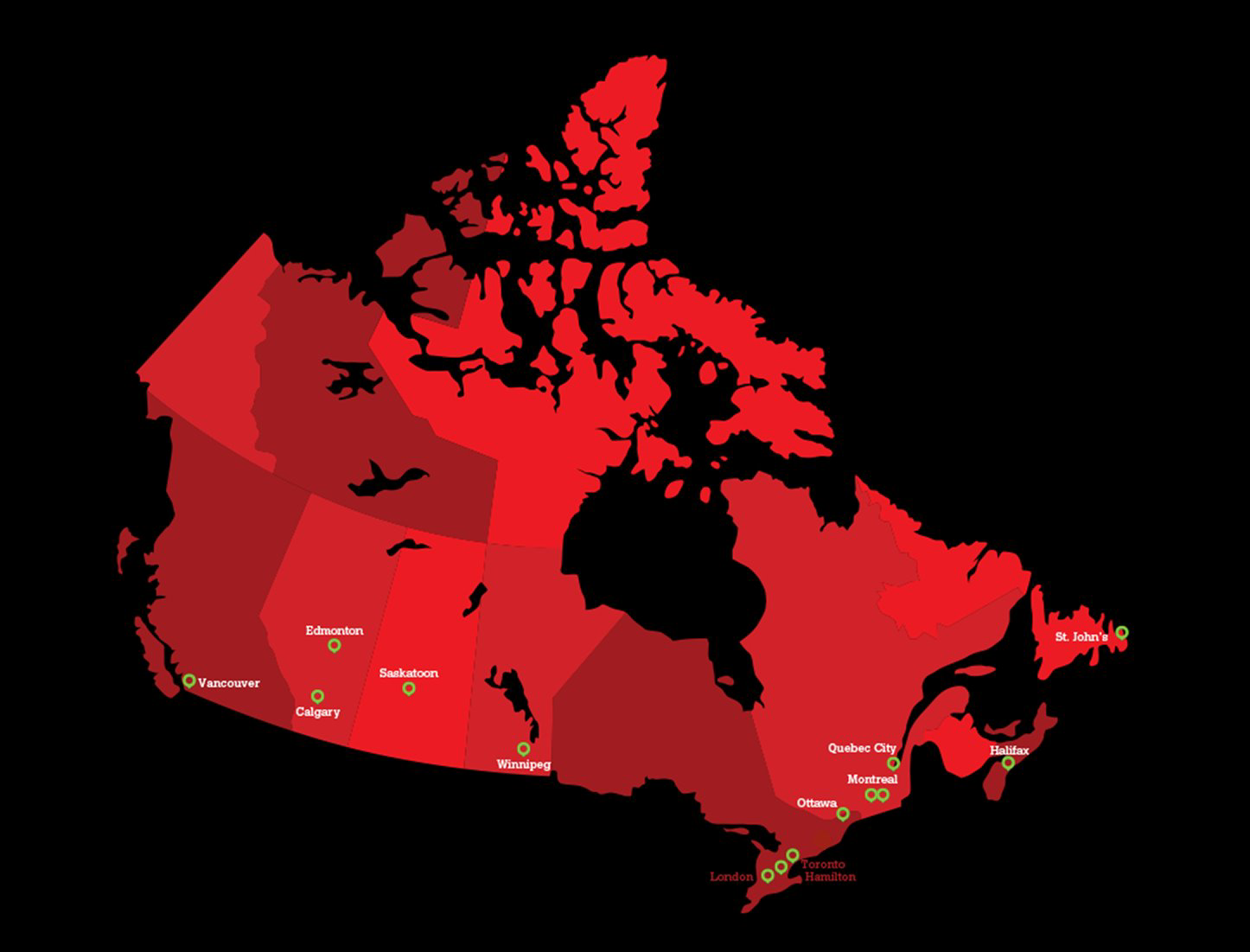

**eTable 2. Proportion of children with unavailable vaccination data.**

| **Provinces/regions of resident** | **Total** | **Unavailable, n** | **Unavailable %** |
| --- | --- | --- | --- |
| Non-Canadian Resident | 73 | 30 | 41.1 |
| Saskatchewan | 2003 | 803 | 40.1 |
| Northwest Territories | 83 | 27 | 32.5 |
| Alberta | 28909 | 8828 | 30.5 |
| Nunavut | 224 | 64 | 28.6 |
| Prince Edward Island | 21 | 4 | 19.0 |
| Ontario | 15599 | 2791 | 17.9 |
| Yukon | 18 | 1 | 5.6 |
| New Brunswick | 82 | 4 | 4.9 |
| British Columbia | 5924 | 174 | 2.9 |
| Quebec | 15386 | 240 | 1.6 |
| Manitoba | 2303 | 25 | 1.1 |
| Newfoundland and Labrador | 2402 | 8 | 0.3 |
| Nova Scotia | 1901 | 3 | 0.2 |
| NA | 454 | 8 | 1.8 |

NA = not available

### **eTable 3. Comparison of baseline characteristics between included and those excluded due to missing vaccination status, 2024-2025 season.**

| **Variable, no. (%)** | **Included (n=17316)** | **Excluded (n=4749)** | **P-value** |
| --- | --- | --- | --- |
| Age (months) (median [IQR]) | 41.0 [18.5, 82.3] | 99.7 [61.5, 150.5] | <0.001 |
| Age group |  |  | <0.001 |
| 6 to <24 months | 5742 (33.2) | 580 (12.2) |  |
| 24 to <60 months | 5228 (30.2) | 566 (11.9) |  |
| 5 to <10 years | 3866 (22.3) | 1743 (36.7) |  |
| 10 to <18 years | 2480 (14.3) | 1860 (39.2) |  |
| Sex |  |  | 0.67 |
| Female | 7963 (46.0) | 2144 (45.1) |  |
| Male | 9350 (54.0) | 2604 (54.8) |  |
| Other | 3 (0.07) | 1 (0.02) |  |
| Median total household income quintile (CAD) |  |  | <0.001 |
| $40,800–71,500 | 3484 (20.2) | 475 (10.1) |  |
| $71,501–83,000 | 3535 (20.5) | 773 (16.4) |  |
| $83,001–94,000 | 3435 (19.9) | 880 (18.7) |  |
| $94,001–110,000 | 3558 (20.6) | 1138 (24.1) |  |
| $110,001–240,000 | 3222 (18.7) | 1447 (30.7) |  |
| Missing | 82 | 36 |  |
| Province/territory of residence |  |  | <0.001 |
| Alberta | 4016 (23.2) | 2658 (56.0) |  |
| British Columbia | 2798 (16.2) | 114 (2.4) |  |
| Manitoba | 653 (3.8) | 10 (0.2) |  |
| Newfoundland and Labrador | 844 (4.9) | 1 (0.0) |  |
| Nova Scotia | 611 (3.5) | 0 (0.0) |  |
| Ontario | 3935 (22.7) | 1428 (30.1) |  |
| Others combined | 63 (0.4) | 32 (0.7) |  |
| Quebec | 4216 (24.4) | 107 (2.3) |  |
| Saskatchewan | 173 (1.0) | 397 (8.4) |  |
| Missing | 7 | 2 |  |
| Diagnosed comorbidity | 5766 (33.3) | 1465 (30.8) | 0.002 |
| Setting |  |  | <0.001 |
| ED visit only | 10165 (58.7) | 2882 (60.7) |  |
| Hospitalized (not ICU) | 6167 (35.6) | 1556 (32.8) |  |
| ICU | 984 (5.7) | 311 (6.5) |  |
| Influenza A | 3700 (21.4) | 924 (19.5) | 0.006 |
| Influenza B | 1074 (6.2) | 266 (5.6) | 0.15 |
| Month of test |  |  | <0.001 |
| January | 3274 (18.9) | 858 (18.1) |  |
| February | 3320 (19.2) | 818 (17.2) |  |
| March | 2323 (13.4) | 701 (14.8) |  |
| April | 1173 (6.8) | 482 (10.1) |  |
| May | 705 (4.1) | 365 (7.7) |  |
| June | 132 (0.8) | 61 (1.3) |  |
| October | 101 (0.6) | 14 (0.3) |  |
| November | 2406 (13.9) | 509 (10.7) |  |
| December | 3882 (22.4) | 941 (19.8) |  |

### **eTable 4. Comparison of baseline characteristics between included children and those excluded due to missing vaccination status, 2025-2026 season.**

| **Variable, no. (%)** | **Included (n=13417)** | **Excluded (n=4152)** | **P-value** |
| --- | --- | --- | --- |
| Age (months) (median [IQR]) | 33.5 [16.7, 74.8] | 99.98 [64.5, 154.5] | <0.001 |
| Age group |  |  | <0.001 |
| 6 to <24 months | 5197 (38.7) | 460 (11.1) |  |
| 24 to <60 months | 4063 (30.3) | 458 (11.0) |  |
| 5 to <10 years | 2318 (17.3) | 1600 (38.5) |  |
| 10 to <18 years | 1839 (13.7) | 1634 (39.4) |  |
| Sex |  |  | 0.23 |
| Female | 6158 (45.9) | 1951 (47.0) |  |
| Male | 7258 (54.1) | 2201 (53.0) |  |
| Missing | 1 | 0 |  |
| Median total household income quintile (CAD) |  |  | <0.001 |
| $36,400–71,000 | 2730 (20.4) | 402 (9.7) |  |
| $71,001–83,000 | 2628 (19.7) | 698 (16.9) |  |
| $83,001–96,000 | 2953 (22.1) | 931 (22.6) |  |
| $96,001–110,000 | 2433 (18.2) | 931 (22.6) |  |
| $110,001–240,000 | 2623 (19.6) | 1166 (28.2) |  |
| Missing | 50 | 24 |  |
| Province/territory of residence |  |  | <0.001 |
| Alberta | 3625 (27.0) | 2553 (61.5) |  |
| British Columbia | 917 (6.8) | 19 (0.5) |  |
| Manitoba | 505 (3.8) | 4 (0.1) |  |
| Newfoundland and Labrador | 773 (5.8) | 3 (0.1) |  |
| Nova Scotia | 537 (4.0) | 1 (0.0) |  |
| Ontario | 3292 (24.5) | 1162 (28.0) |  |
| Others combined | 72 (0.5) | 32 (0.8) |  |
| Quebec | 3403 (25.4) | 101 (2.4) |  |
| Saskatchewan | 291 (2.2) | 275 (6.6) |  |
| Missing | 2 | 2 |  |
| Diagnosed comorbidity | 4554 (33.9) | 1345 (32.4) (n=4151) | 0.08 |
| Setting |  |  | <0.001 |
| ED visit only | 7691 (57.3) | 2618 (63.1) |  |
| Hospitalized (not ICU) | 4822 (35.9) | 1320 (31.8) |  |
| ICU | 904 (6.7) | 214 (5.2) |  |
| Influenza A | 3560 (26.5) | 971 (23.4) | <0.001 |
| Influenza B | 1243 (9.3) | 562 (13.5) | <0.001 |
| Month of test |  |  | <0.001 |
| January | 2039 (15.2) | 545 (13.1) |  |
| February | 1519 (11.3) | 473 (11.4) |  |
| March | 1552 (11.6) | 583 (14.0) |  |
| April | 1344 (10.0) | 441 (10.6) |  |
| May | 881 (6.6) | 288 (6.9) |  |
| June | 217 (1.6) | 103 (2.5) |  |
| November | 1299 (9.7) | 396 (9.5) |  |
| December | 4566 (34.0) | 1323 (31.9) |  |

**eTable 5. Proportions of vaccinated children with vaccination status determined by provincial immunization registries, medical records, and caregiver report, by influenza season and province.**

|  | **Total** | **Immunization registries** | **Medical records** | **Caregiver report** |
| --- | --- | --- | --- | --- |
| **2024-2025** |  |  |  |  |
| Alberta | 978 | 971 (99.3%) | 7 (0.7%) | 0 (0.0%) |
| British Columbia | 472 | 463 (98.1%) | 0 (0.0%) | 9 (1.9%) |
| Manitoba | 89 | 81 (91.0%) | 8 (9.0%) | 0 (0.0%) |
| Newfoundland and Labrador | 81 | 80 (98.8%) | 1 (1.2%) | 0 (0.0%) |
| Nova Scotia | 138 | 137 (99.3%) | 1 (0.7%) | 0 (0.0%) |
| Ontario | 484 | 1 (1.5%) | 112 (23.1%) | 371 (76.7%) |
| Quebec | 333 | 330 (99.1%) | 3 (0.9%) | 0 (0.0%) |
| Saskatchewan | 6 | 0 (0.0%) | 1 (16.7%) | 5 (83.3%) |
| **2025-2026** |  |  |  |  |
| Alberta | 891 | 883 (99.1%) | 7 (0.8%) | 1 (0.1%) |
| British Columbia | 172 | 169 (98.3%) | 0 (0.0%) | 3 (1.7%) |
| Manitoba | 71 | 71 (100.0%) | 0 (0.0%) | 0 (0.0%) |
| Newfoundland and Labrador | 149 | 149 (100.0%) | 0 (0.0%) | 0 (0.0%) |
| Nova Scotia | 120 | 120 (100.0%) | 0 (0.0%) | 0 (0.0%) |
| Ontario | 344 | 0 (0.0%) | 147 (42.7%) | 197 (57.3%) |
| Quebec | 296 | 294 (99.3%) | 0 (0.0%) | 2 (0.7%) |
| Saskatchewan | 5 | 0 (0.0%) | 1 (20.0%) | 4 (80.0%) |

**eTable 6. Sensitivity analyses of vaccine effectiveness (VE) against any influenza in any setting for the 2024-2025 and 2025-2026 seasons.**

|  | **N** | **VE** |
| --- | --- | --- |
| **2024-2025** |  |  |
| Primary analysis | 17316 | 47% (40%–53%) |
| Sensitivity analysis excluding provinces with >20% of children with unavailable vaccination data^1^ | 13077 | 47% (39%–54%) |
| Sensitivity analysis excluding provinces with >10% of children with unavailable vaccination data^2^ | 9139 | 48% (38%–56%) |
| Sensitivity analysis re-classifying caregiver-report-only children as unvaccinated | 17316 | 48% (41%–55%) |
| Sensitivity analysis excluding caregiver-report-only children | 16927 | 48% (41%–55%) |
| **2025-2026** |  |  |
| Primary analysis | 13417 | 53% (46%–59%) |
| Sensitivity analysis excluding provinces with >20% of children with unavailable vaccination data^1^ | 9447 | 45% (36%–53%) |
| Sensitivity analysis excluding provinces with >10% of children with unavailable vaccination data^2^ | 6151 | 45% (35%–54%) |
| Sensitivity analysis re-classifying caregiver-report-only children as unvaccinated | 13417 | 55% (49%–61%) |
| Sensitivity analysis excluding caregiver-report-only children | 13210 | 55% (49%–61%) |

^1^Non-Canadian Resident, Saskatchewan, Northwest Territories, Alberta, Nunavut

^2^Non-Canadian Resident, Saskatchewan, Northwest Territories, Alberta, Nunavut, Prince Edward Island, Ontario
